# A CRISPR mediated point-of-care assay for the detection of mucosal calprotectin in an animal model of ulcerative colitis

**DOI:** 10.1101/2024.03.23.24304787

**Authors:** Selena Chia, Tianruo Guo, Ewa M. Goldys, Sophie C. Payne, Nigel H. Lovell, Mohit N. Shivdasani, Fei Deng

## Abstract

Inflammatory bowel disease (IBD) is a chronic disorder associated with inflammation in the gastrointestinal tract, leading to a range of debilitating symptoms. Fecal calprotectin is an established biomarker for ulcerative colitis (UC), one of the main IBD diseases, which provides indications of the presence and severity of inflammation in the digestive tract. Enzyme-Linked Immunosorbent Assay (ELISA) as a gold standard approach for fecal calprotectin detection is time-consuming and impractical in point-of-care settings. Moreover, obtaining fecal samples from patients is challenging and inhibits longitudinal monitoring. To overcome these limitations, we designed a new approach for detecting calprotectin which leverages clustered regularly interspaced short palindromic repeats (CRISPR)/Cas technology. We successfully developed a portable tube-based CRISPR/Cas assay for point-of-care testing of calprotectin. This assay showed a detection range from 1-10000 ng/mL (over 4 log units), using both fluorescent and colorimetric analytical techniques. The established assay was further validated through measurements in mucosal samples obtained in an anesthetised preclinical rodent model of UC, with 2-3 times higher calprotectin concentration detected in UC rat samples compared to that of healthy control animals. This point-of-care test may provide a rapid, precise, and user-friendly approach for the diagnosis and monitoring of IBD through mucosal sample testing.

## 1. Introduction

Inflammatory bowel disease (IBD) is a chronic debilitating disorder characterized by the inflammation in the gastrointestinal tract^1^ and encompasses two main types of this incurable condition, Crohn’s disease (CD) and Ulcerative Colitis (UC)^2^. IBD can present symptoms such as diarrhea, bloody stools, abdominal pain, and fatigue due to impaired digestive function, nutrient absorption and waste elimination^3^. Effective IBD diagnosis methods include endoscopy/biopsy, and biomarker detection combined with multiparameter clinical scoring based on presented symptoms. Notably, biomarker detection provides a sensitive and non-invasive add-on method to the traditionally invasive and subjective diagnostic methods for IBD^4^, and in many cases can alone have positive predictive power in tracking disease progression. Calprotectin (CP) in feces has emerged as a promising biomarker for IBD diagnostics particularly in UC, with excellent sensitivity (92.6%) and specificity (88.9%), allowing clinicians to not only distinguish CD from UC as levels of fecal CP are much lower in CD, but also distinguish IBD from other conditions such as irritable bowel syndrome^5^. Previous studies show that fecal calprotectin concentrations are well correlated with disease activity assessed through clinical symptoms, endoscopic findings and histological markers^6, 7^. This correlation indicates the potential for exploring the clinical utility of CP in other bodily fluids that are less convenient to sample than fecal sampling, such as blood^8^. Indeed, there have been several studies^9, 10^ that have attempted measurements of CP in serum and shown good correlation of these measurements to not only fecal CP, but also other clinical outcomes. Overall, this highlights the significance of CP detection as a sensitive, specific and less-invasive method of IBD diagnosis and monitoring^11^.

To date, diverse bioanalytical assays have been utilized for calprotectin detection, including Enzyme-Linked Immunosorbent Assay (ELISA), turbidimetric assays, lateral flow assays and chemiluminescent immunoassays. However, these assays are limited by a number of factors, including the high cost and instability associated with the use of antibodies^12^. Moreover, ELISA test processing requires specialized technicians with centralized equipment, making it tedious and labor intensive, resulting in extended assay times^13^. While turbidimetric and lateral flow assays offer simplicity and rapid results, the methods are limited by their low dynamic range and sensitivity^14^. Finally, fully automated quantitative assays such as the Diasorin LIAISON CP require large and expensive instrumentation^15^. Importantly, none of the existing measurement techniques are viable for point-of-care testing (POCT). A recent study^16^ escribed the development and validation of a novel flow immune-chromatography assay for measuring fecal CP, which could be used as a POCT or at-home test. However, the POCT assay described in this study (Preventis SmarTest Calprotectin Home, Preventis, Bensheim, Germany) is only suitable for measuring fecal CP levels, highlighting the clear unmet need for a point-of-care CP detection method that offers high specificity, high sensitivity, and accessibility for other biological samples.

Clustered Regularly Interspaced Short Palindromic Repeats (CRISPR) Cas12a protein originates from the bacterial immune system and has been widely applied in molecular diagnostic technologies^17^. Its highly sequence-specific target recognition and exceptional *trans*-cleavage activity make it an effective nucleic acid diagnosis tool ^18^. For the application of CRISPR/Cas12a technology for non-nucleic acid detection, the non-nucleic acid input must be converted into nucleic acid signals to activate the CRISPR/Cas system, which has been demonstrated using aptamers ^19^, antibody-ssDNA ^20-22^, antibody-Cas conjugate ^23^ etc., with the limit of detection of small proteins, such as cytokines, ranging from 1 pg/ml to 1 fg/mL ^20, 21^. Given CP is also a small protein composed of two calcium binding proteins, CRISPR/Cas12a holds significant potential for ultrasensitive detection of CP. Additionally, Cas12a sensors can be utilized in portable assays, such as paper-based devices, suitable for POCT ^24, 25^.

In this study, we present two CRISPR/Cas assays for the detection of CP. A fluorescent CRISPR/Cas12a assay was first established in a tube-based format, which exhibited exceptional performance with a limit of detection (LOD) of 1 ng/mL. Subsequently, we built a colorimetrical CRISPR/Cas12a assay suitable for a point-of-care setting, with the same LOD (1 ng/mL). Both assays were validated through measurements from mucosal samples obtained from a preclinical rodent UC model based on 2,4,6-trinitrobenzene sulphonic acid (TNBS) ^26^ injections. Following an intra-rectal injection, TNBS binds to proteins inside the colon and forms a complex that is recognized by the immune system. This causes a sudden but localized inflammatory reaction at the site of TNBS administration. Mucosal samples were specifically chosen, as these would be easy to obtain in a POCT setting and could be obtained at the same time as an endoscopic examination^27^. The mucosal samples from IBD rats were compared to samples obtained from healthy rats to demonstrate the future clinical suitability of our CRISPR technique.

## 2. Material and Methods

### 2.1 Materials

The materials for this study included: EnGen® Lba Cas12a (Cpf1) protein (New England Biolab), Sodium chloride (Sigma), Tris-hydrochloride (Sigma), Manganese(II) sulfate (MnSO_4_, Sigma), 2,4,6-trinitrobenzene sulphonic acid (TNBS), Bovine Serum Albumin (Sigma), DNase/RNase free water (ThermoFisher), recombinant calprotectin (R&D, 8226-S8), phosphate buffered saline (PBS) (Sigma, 10 mM, pH=7.4), rat CP ELISA kit (EKC38778-96T, Biomatik) and HybriDetect – Universal Lateral Flow Assay Kit (Milenia biotec).

All DNA and RNA oligos (Table 1) were synthesized and modified by Sangon Bio-Tech Ltd.

**Table 1.**
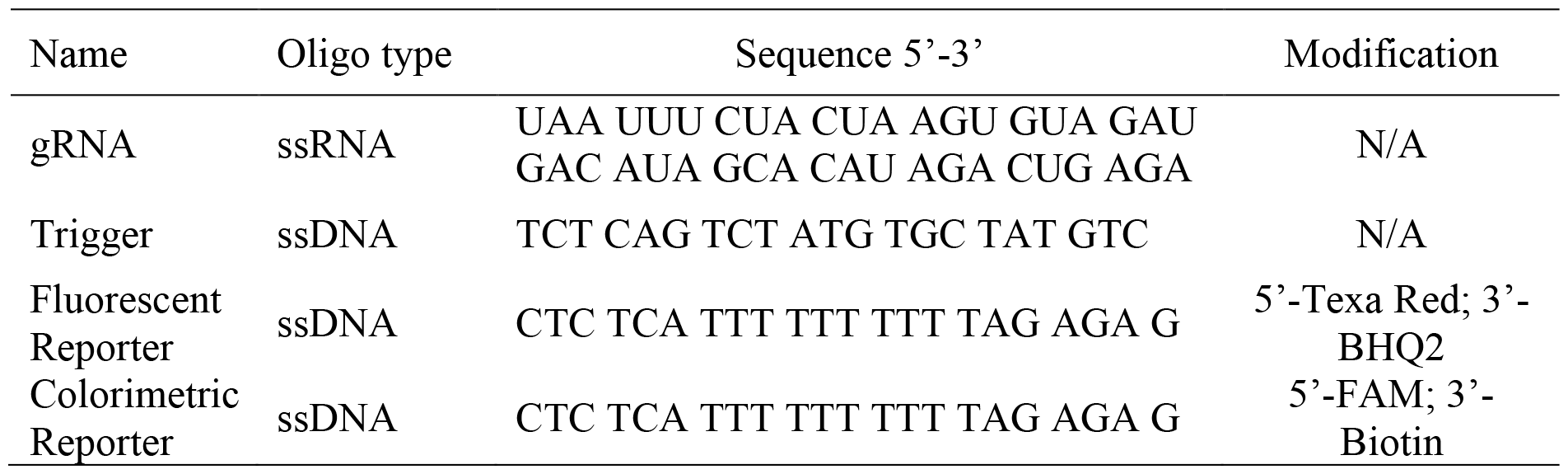
DNA and RNA oligos used in this study.

### 2.2 Optimization of the CRISPR/Cas12a biosensing system

The CRISPR/Cas12a reaction buffer was first prepared by combining the following reagents: 50 mM NaCl, 10 mM Tris-HCl, 10 μM MnSO_4_, 100 μg/mL BSA, pH 7.9@25°C.

Following this, the CRISPR/Cas12a reaction solution was prepared. In brief, 1 μL of 100 μM of Cas12a protein, 5 μL of 20 μM of guide RNA (gRNA), and 6 μL of 100 μM of fluorescent reporter were added into 3.6 mL of reaction buffer. The prepared solution was stored at 4 °C before use. Prior to testing with recombinant CP and rat mucosa samples, a number of optimization experiments were conducted. All fluorescence signals were tested using a SpectraMax iD5 multi-Mode Microplate Reader (λex: 570 nm; λem: 615 nm).

#### Optimization of manganese ion concentration

Mn^2+^ is an essential component to stabilize the structure of Cas12a protein^28^. To optimize the concentration of Mn^2+^, different concentrations of Mn^2+^ (0, 1.25, 2.5, 5, 10, 20, 30 and 40 μM) based CRISPR/Cas12a reaction buffers were prepared. Subsequently, CRISPR/Cas12a reaction solution was prepared. Following this step, 40 nM of trigger DNA was added to trigger the CRISPR/Cas12a reaction and incubated at room temperature for two hours, then tested.

#### Optimization of trigger DNA concentration

To optimize the trigger DNA concentration, different concentrations of trigger artificial singled-stranded DNA (ssDNA) (0, 2.5, 5, 10, 20, 40, 60, 80 and 100 nM) were added to 100 μL of CRISPR/Cas12a reaction solution and incubated at room temperature for two hours, then tested.

#### Optimization of temperature

To investigate the optimum reaction temperature, 40 nM of trigger DNA was added to 100 μL of CRISPR/Cas12a reaction solution and incubated at room temperature or 37 °C for two hours, then tested.

#### Optimization of dithiothreitol

In our previous research, dithiothreitol (DTT) was demonstrated to be an effective chemical enhancer to Cas12a ribonucleoprotein (RNP) ^18^. In this study, different concentrations (0 and 10mM) of DTT were added to the CRISPR/Cas12a reaction solution (3.6 mL). Subsequently, 40 nM of trigger DNA was added to trigger the CRISPR/Cas12a reaction, and the solution was tested.

### 2.3 Evaluation of the biosensing performance

Following optimization of several components, we then prepared an optimized CRISPR/Cas12a reaction buffer by combining the following reagents: 50 mM NaCl, 10 mM Tris-HCl, 10 μM MnSO_4_, 100 μg/mL BSA, 10mM DTT, pH 7.9@25°C.

Subsequently, we prepared an optimized CRISPR/Cas12a reaction solution using 1 μL of 100 μM of Cas12a protein, 5 μL of 20 μM of guide RNA (gRNA), and 6 μL of 100 μM of fluorescent reporter added to 3.6 mL of the optimized CRISPR/Cas12a reaction buffer. The prepared solution was stored at 4 °C before use.

#### Evaluation of sensitivity

To investigate the sensitivity of the optimized CRISPR/Cas12a biosensing system for the detection of CP, different concentrations of recombinant CP (0, 1 ng/mL, 10 ng/mL, 100 ng/mL, 1 μg/mL and 10 μg/mL, reconstituted in PBS) were added to 100 μL of the optimized CRISPR/Cas12a reaction solution. Subsequently, 40 nM of trigger DNA was added into 100 μL of prepared CRISPR/Cas12a reaction solution and incubated at room temperature for one hour, then tested as room temperature was found to be more optimal than at 37 °C.

#### Evaluation of specificity

To investigate the specificity of the optimized CRISPR/Cas12a biosensing system, a variety of recombinant interference proteins were added to 100 μL of the optimized CRISPR/Cas12a reaction solution, including Interferon-γ (IFN-γ, 10 μg/mL), Interleukin-6 (IL-6, 10 μg/mL), Interleukin-10 (IL-10, 10 μg/mL), Tumor Necrosis Factor-α (TNF-α, 10 μg/mL), Interleukin-1β (IL-1β, 10 μg/mL) and BSA protein (10 mg/mL). CP as the target protein was also tested at a concentration of 10 μg/mL. Subsequently, 40 nM of trigger DNA was added, and the solution incubated at room temperature for one hour, then tested.

#### Evaluation of stability

To investigate the stability of the optimized CRISPR/Cas12a biosensing system, the prepared CRISPR/Cas12a solution was stored at 4 °C for four weeks. The biosensing performance was evaluated every week. For each test, 100 μL of CRISPR/Cas12a storage solution was taken out, and CP was added to make a final concentration of 10 μg/mL of CP. Following this, 40 nM of trigger DNA was added to 100 μL of the prepared CRISPR/Cas12a reaction solution and incubated at room temperature for one hour, then tested.

#### Evaluation of reproducibility

To investigate the reproducibility of the optimized CRISPR/Cas12a biosensing system, the CRISPR/Cas12a solution and CP testing protocols were repeated eight times in total. In brief, the optimized CRISPR/Cas12a reaction solution was prepared. Subsequently, CP was added to the CRISPR reaction solution resulting in a final concentration of 10 μg/mL of CP. Following this, 40 nM of trigger DNA was added to 100 μL of the prepared CRISPR/Cas12a reaction solution and incubated at room temperature for one hour, then tested.

### 2.4 In vivo rat model of acute IBD and intestinal mucosa sample collection

All procedures were reviewed and approved by the UNSW Animal Care and Ethics committee (No. 21/130A) and carried out in compliance with the Australian Code for the Care and Use of Animals for Scientific Purposes (8th Edition 2013, National Health and Medical Research Council of Australia) and the Prevention of Cruelty to Animals Act (1986). Male Long Evans rats (age 15-20 weeks, Animal Resource Centre, Western Australia) were fasted for 12 hours and anaesthetized (2-3% vaporized isoflurane in 2 L/min oxygen). Buprenorphine (0.02 mg/kg, Cenvet) was injected subcutaneously as a preemptive analgesic.

As described previously^29, 30^, acute colitis was induced by injecting 0.25 ml of 1% TNBS (in 50% ethanol, Sigma) 8 cm into the colon using a custom catheter^30^. Intestinal mucosa was collected hourly over 4-6 hours using filter paper technology validated in humans, starting from 30 minutes post-injection of TNBS ^31^. For each test, a piece of filter paper (Whatman 42.5 mm) wrapped around a custom tube was inserted ∼8 cm into the colon, incubated for 20 minutes, then removed and transferred to a tube containing 1 mL Tris buffer solution (0.1 M Tris, 0.3% human serum albumin, 0.01% sodium azide, and 0.002% Tween)^27^. Following several measurements, animals were euthanised with Sodium Pentobarbital. The concentration of CP released in mucosal sample solutions were then measured using our CRISPR assays and compared to a commercial ELISA kit (DS8900, R&D). We chose to conduct mucosal measurements of CP as opposed to fecal or blood samples, as collecting mucosal samples from anesthetized rats was convenient, has been established in humans and has been shown to contain concentrations of other biomarkers that can clearly differentiate between active and inactive UC in human IBD patients as well as differences between IBD patients and healthy humans. In addition, mucosal samples would be relatively easier to obtain in a POCT setting, compared to blood or stool samples, and could be obtained at the same time as an endoscopic examination^27^.

### 2.5 Application of fluorescent CRISPR/Cas12a biosensor for the detection of CP from rat mucosa samples

For the fluorescent CRISPR/Cas12a assay measurements, the optimized CRISPR/Cas12a reaction solution was first prepared by using 1 μL of 100 μM of Cas12a protein, 5 μL of 20 μM of guide RNA (gRNA), and 6 μL of 100 μM of fluorescent reporter added to 3.6 mL of the standard CRISPR/Cas12a reaction buffer. The prepared solution was stored at 4 °C before use.

Following this, 10 μL of the mucosal sample solution and 40 nM of trigger DNA were added into 90 μL of the optimized CRISPR/Cas12a reaction solution and incubated at room temperature for one hour, then tested.

### 2.6 Application of Lateral Flow Assay assisted CRISPR/Cas12a biosensor for the detection of CP from rat mucosal samples

As the fluorescent based assay required the use of a microplate reader for quantitative measurements, we also aimed in this study to develop a more point-of-care viable test for CP detection. For this, we developed a colorimetric assay which made the use of lateral flow assay (LFA) technology in the form of test strips. To do this, a colorimetric CRISPR/Cas12a reaction solution was first prepared by using 1 μL of 100 μM of Cas12a protein, 5 μL of 20 μM of guide RNA (gRNA), and 6 μL of 100 μM of colorimetric reporter added to 3.6 mL of the optimized CRISPR/Cas12a reaction buffer. The prepared solution was stored at 4 °C before use.

Following this, 10 μL of the 1000 times diluted mucosal sample solution and 40 nM of trigger DNA were added into 90 μL of the optimized CRISPR/Cas12a reaction solution and incubated at room temperature for one hour, then tested. Subsequently, 6 μL of the reaction mixture was added to 94 μL of HybriDetect assay buffer (Milenia) and run on HybriDetect lateral flow strips (Milenia). The colorimetric test results were quantified using the Axxin LFA reader.

### 2.7 Histology of in vivo rat model colonic tissue

At 4 hours post-TNBS, rats were euthanized (350 mg/kg Lethabarb, intracardial injection) and inflamed colon adjacent to TNBS injection location was removed. For the control group, colonic tissue was from normal rats at 5 cm and 8 cm into the colon. As described previously ^32^, colon tissues were placed into cold PBS/Nicardipine (0.15 M NaCl in 0.01 M sodium phosphate buffer, pH 7.2, 1 μM Nicardipine), cut longitudinally and pinned out onto a balsa board with mucosa side up and the colon wall in contact with the balsa board. The intraluminal content was gently removed by washing with cold PBS. The tissue was fixed in Zamboni’s (2% formaldehyde plus 0.2% picric acid in 0.1 M sodium phosphate buffer, pH 7.4) overnight and placed in 70% ethanol prior to histological processing. Samples were processed in Leica HistoCore PEARL and embedded in Paraffin, sectioned 4 μm on Leica RM2165 microtome and stained with haematoxylin and eosin (H&E). Sections were mounted with dibutylphthalate polystyrene xylene (DPX) and imaged on Olympus VS200 using 20× NA 0.8 objective. A histopathologist, blinded to experimental conditions, used H&E-stained sections to confirm TNBS-induced histopathological damage in the tissue.

### 2.8 Statistical Methods

Statistical differences or their absence (indicated by “ns”) were assessed by ANOVAs with multiple comparisons or two-tailed t-tests and stated where appropriate, with stars indicating *p*-values (* *p*<0.05; ** *p*<0.01; *** *p*<0.005).

## 3 Results

### 3.1 Schematic of CRISPR/Cas12a biosensor for the detection of CP

The CRISPR/Cas12a biosensing system consists of Cas12a RNP and fluorescent quenched nucleic acid reporters. With the addition of artificial singled-stranded DNA (ssDNA) acting as a trigger, Cas12a RNP will be activated to cleave the fluorescent reporters, leading to a certain amount of fluorescence signal. With the introduction of CP into the biosensing system, a higher fluorescence signal readout was observed (Fig. 1 shows one example dataset from our experiments). This indicates an enhancement effect of CP on the CRISPR/Cas12a biosensing system^33^. In addition, a positive correlation between the CP concentration and fluorescence intensity was observed, which provided a quantification approach for the testing of CP from biological samples. This CRISPR/Cas12a biosensing system used a single pot reaction so it is suitable for a tube-based portable assay.

**Figure 1:**
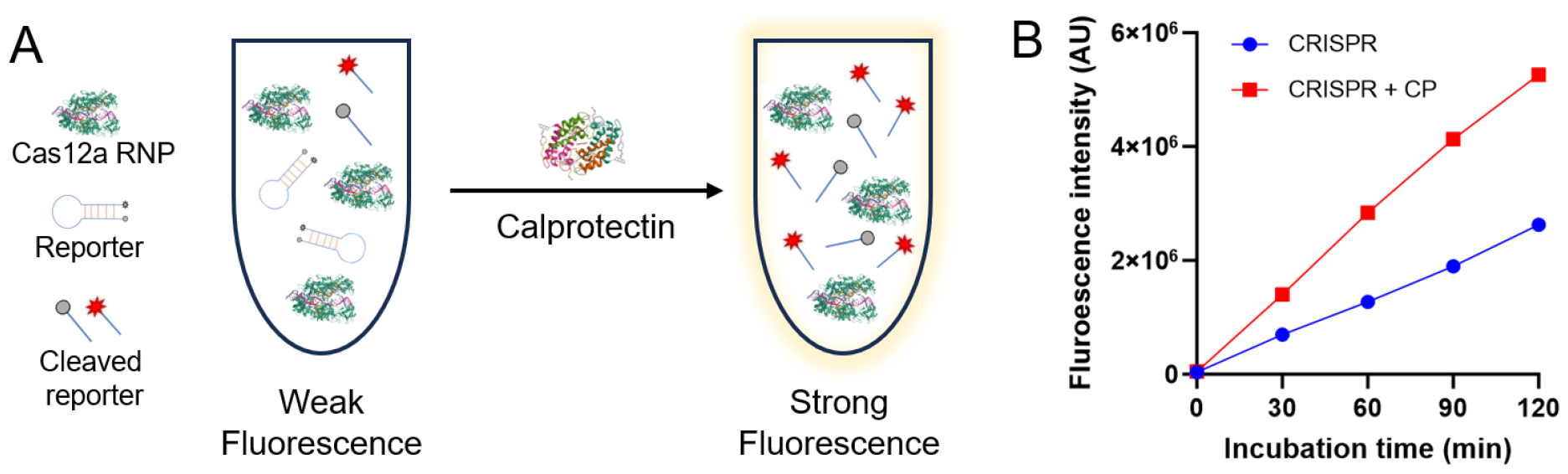
Schematic figure of CRISPR/Cas12a biosensing system for calprotectin detection. (A) CP enhances the biosensing performance of CRISPR/Cas12a biosensing system for artificial triggers; (B) Higher fluorescence signal was observed in CP-enhanced CRISPR/Cas12a biosensing system (example dataset from one of our experiments).

### 3.2 Optimization of the key parameters of CRISPR/Cas12a biosensor

In the optimization phase, we sought to determine the concentration of key parameters to elicit the strongest fluorescence. Varying the concentration of the trigger ssDNA led to an increased signal which saturated at a concentration of 40 nM (one-way ANOVA, <u>Fig. 2</u>A). When optimizing temperature (Fig. 2B), we found no statistical significance (two-way ANOVA, *p>0*.*05*) in the fluorescence intensity between room temperature and 37°C at 60 minutes, however, there was a significant effect of temperature for an incubation time of two hours (*p*<0.005). Subsequently, a significant (two-way ANOVA, p<0.0001) enhancement effect of DTT on fluorescence was observed at 120 min (Fig. 2C), where the signal increased by a factor of 3.9 compared to without DTT. DTT enhancement was also significant for a 60 min incubation (p<0.0001). Finally, we investigated the concentration of manganese ion (one-way ANOVA, Fig. 2D) and found 10 μM to be the optimum concentration. These optimized values were utilized in subsequent experiments involving detection of recombinant CP and CP present in rat mucosal solution samples.

**Figure 2:**
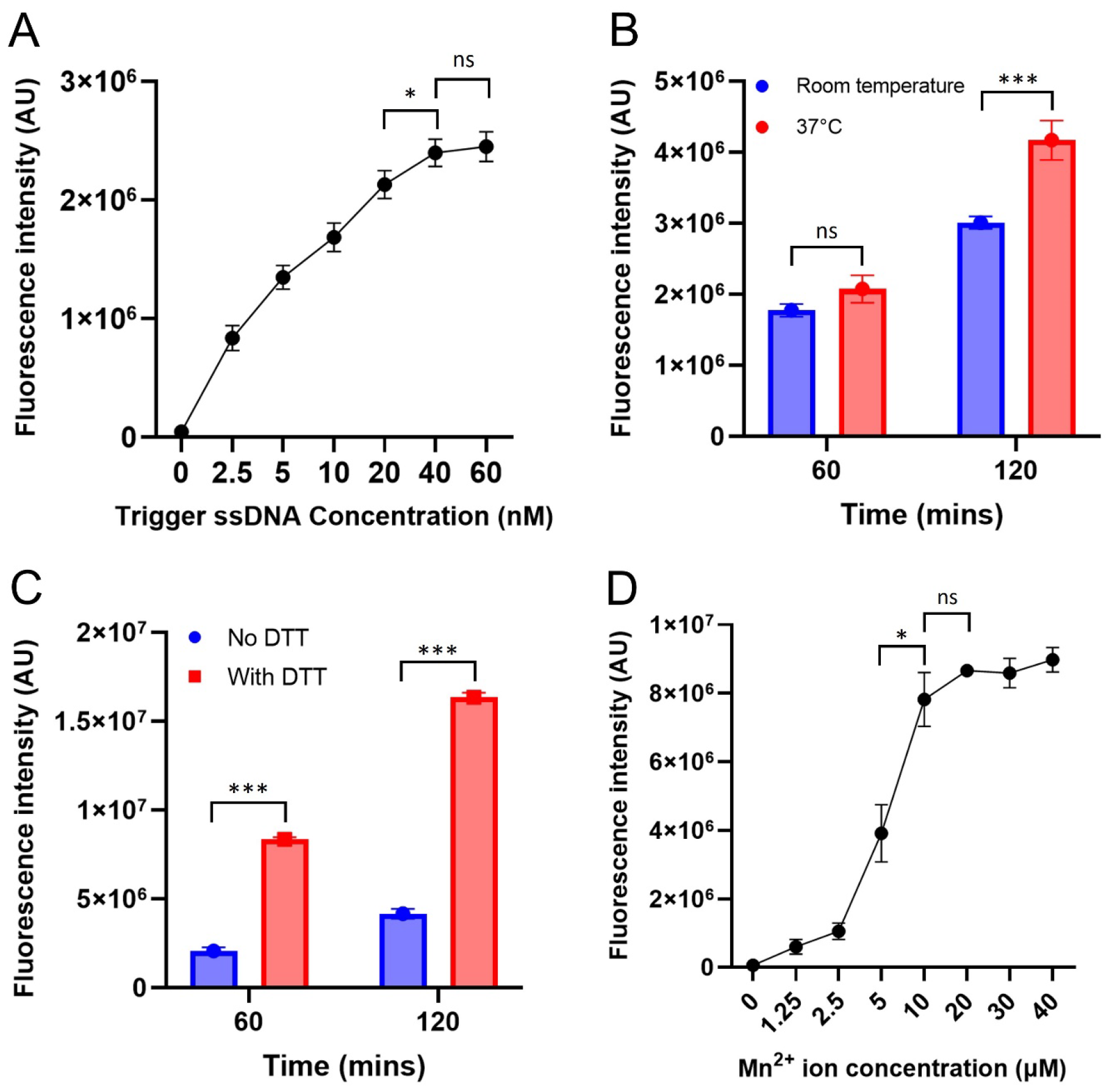
Optimisation of CRISPR/Cas12a biosensor components to realize the optimum biosensing performance. (A) Fluorescence intensity measured from CRISPR-target activity with various trigger ssDNA concentrations at 60 mins (n=3). One-way ANOVA multiple comparisons: 40 nM vs 60 nM was not significant, 40nM vs the rest was significant (P<0.05). (B) Fluorescence from CRISPR-target activity with 40 nM trigger ssDNA concentration over time in room temperature and 37°C temperature (n=3). Two-way ANOVA multiple comparisons: RT 60 min vs 37°C 60 min was not significant (p>0.05), RT 120 min vs 37°C 120 min was significant (p = 0.0014). (C) Presence of DTT in the CRISPR/Cas12a system enhanced the fluorescence signal (n=3). Two-way ANOVA multiple comparisons: 60 min No DTT vs 60 min With DTT was significant (p< 0.0001), 120 min No DTT vs 120 min With DTT was significant (p < 0.0001). (D) The effect of Mn^2+^ ion concentration on the fluorescence signal at 120 minutes (n=3). One-way ANOVA multiple comparisons: 5 μM vs 10 μM was significant (p = 0.0104), 10 μM vs 20 μM was not significant (p > 0.05). (\**p*<0.05; ** *p*<0.01; *** *p*<0.005).

### 3.3 Evaluation of basic biosensor performance

To evaluate the biosensing performance of our established CRISPR/Cas12a biosensor, we investigated its sensitivity, specificity, stability, and reproducibility. As shown in Fig. 3A, the limit of detection was observed to be 1 ng/mL, with an over four orders of magnitude detection range from 1 ng/mL to 10 μg/mL. Our CRISPR assay was also highly specific to detection of calprotectin with the background fluorescence of all the interference proteins measured to be 2.4 × 10^6^, and the intensity measured for calprotectin measured to be 45% greater and significantly different (one-way ANOVA, *p<0*.*0001)* to all other proteins (Fig. 3B). Subsequently, the stability of the biosensor was investigated over a 4-week period using a CP concentration of 10 μg/mL (Fig. 3C), and no difference was observed across weeks, indicating a stable CRISPR/Cas12a mixture over this time. Finally, investigations into the reproducibility (Fig. 3D) found no significant difference (one-way ANOVA) between the fluorescence intensity across the 8 batches tested using a 10 μg/mL concentration of calprotectin.

**Figure 3:**
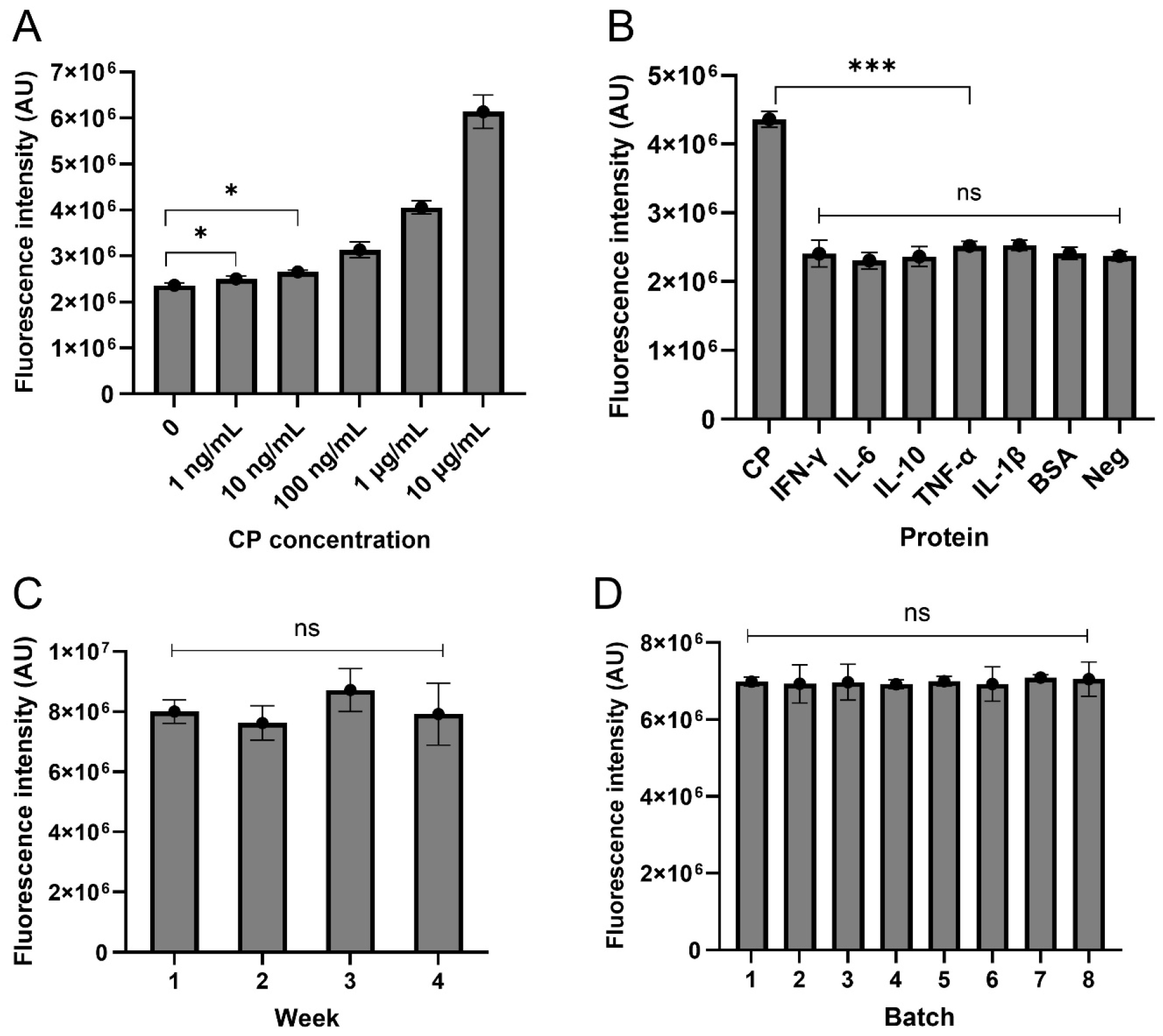
Evaluation of basic biosensor performance. (A) The sensitivity of the biosensor to CP for a 60-min incubation time as measured via fluorescence intensity (n=3). One-way ANOVA multiple comparisons: 0 vs 1 ng/mL was significant (p<0.05). (B) The specificity of the biosensor to a range of proteins for 60-min incubations (n=3). One-way ANOVA multiple comparisons: calprotectin vs each interference protein was significant (p<0.0001). The concentrations of calprotectin, IFN-γ, IL-6, IL-10, TNF-α, IL-1β were 10 μg/mL, the final concentration of BSA was 10 mg/mL, and the control had no targets; (C) The fluorescence intensity of the CRISPR solution did not significantly change over 4 weeks (n=3). One-way ANOVA multiple comparisons, no significant differences were observed among these groups (p>0.05). (D) Fluorescence intensity of 8 batches of CRISPR/Cas solution tested with 10 μg/mL of calprotectin (n=3). One-way ANOVA multiple comparisons, no significant differences were observed among these groups (p>0.05). (\**p*<0.05; ** *p*<0.01; *** *p*<0.005)

### 3.4 Fluorescent CRISPR biosensor for the detection of CP from rat mucosa samples

In normal colonic tissue (n=3), the epithelial layer was intact and undamaged, with a normal distribution of goblet cells and normal crypt architecture (Fig. 4A1). At 4 hours following TNBS injection (n=3), tissue was observed to have bleeding into the mucosal layer, extensive loss of epithelial cells across the majority of the mucosal surface with moderate to severe loss of goblet cells (Fig 4A2). There was a mild infiltration of leucocyte infiltration observed within the mucosal layer. Subsequently, we applied our assay to detect calprotectin level from rat mucosa samples collected from one animal both prior to and after injection of TNBS which induced acute experimental colitis. The fluorescent CRISPR assay was first evaluated (Fig. 4B), which was a plate-based assay requiring instrumentation for fluorescence recording. As shown in Fig. 4C, the fluorescence signal of the diseased samples (n = 3 rats) was significantly different (*p*<0.005) to the controls (n = 3 rats) even for the smallest 30-minute incubation time tested. Based on the established calibration curve in Fig. 3A, the value of CP in control was close to 0, while the value of CP in diseased samples was close to 10 μg/mL. In Figure 4D (incubation times for the sensor kept to 60 mins), three baseline measurements were taken before the induction of TNBS colitis, revealing no noteworthy changes in signal intensity in one rat. Following the injection of TNBS, a notable increase in signal intensity (corresponding to increasing calprotectin concentrations) was observed. A near time-dependent rising trend was noted, suggesting a progressive worsening of the inflammation over time. This result closely matched pre- and post-TNBS serum CP measurements done previously in animal models with experimental colitis^34^.

**Figure 4:**
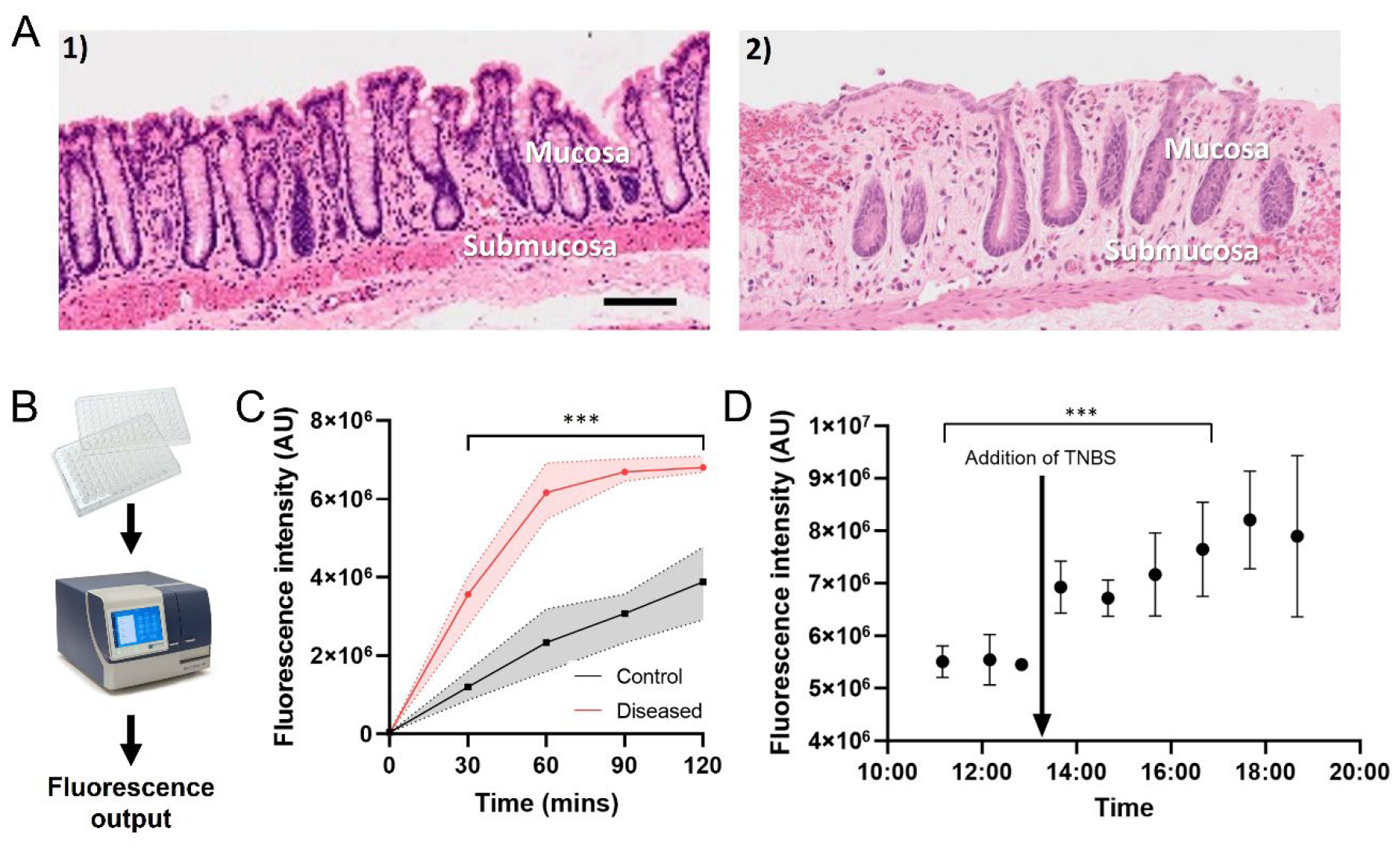
Application of fluorescent CRISPR/Cas12a biosensor for the detection of calprotectin from rat samples. (A) Histological damage following TNBS injection. 1) Representative images of stained sections of the colon from normal tissue. 2) tissue from the inflammatory location (8 cm). Scale bar: 500 μm. (B) Schematic of tube-based assay process: The CRISPR solution is mixed and pipetted into a 96-well plate and the fluorescence is measured using a plate reader to obtain the fluorescence output of the calprotectin level; (C) Fluorescence of rat mucosa samples tested with the optimised CRISPR/Cas12a biosensor (n=3 rats). One-way repeated measures ANOVA with multiple comparisons, significant differences was observed between control and diseased samples (p<0.0001). (D) The application of fluorescent CRISPR/Cas12a biosensor for the longitudinal acute monitoring of CP concentration in mucosa samples in one rat (n= up to 3 repeat measurements at each time point). The timeline is referenced in relation to the surgery time. One-way ANOVA with multiple comparisons, significant differences were observed before and after TNBS (p<0.001). (* *p<*0.05; ** *p*<0.01; *** *p*<0.005; n=3).

### 3.5 LFA assisted CRISPR/Cas12a biosensor for the detection of CP from rat mucosa samples

To realize a point-of-care setting for the CRISPR assay, we integrated the tube-based CRISPR reaction with a lateral flow device (Fig. 5A). The colorimetric calibration curve was first generated (Fig. 5B), with the limit of detection of 1 ng/mL, and detection range from 1 ng/mL to 10 μg/mL. This is visually confirmed in the image of the test strips (Fig. 5B), where the optical density of the test line is darker with increasing CP concentration. Subsequently, we applied the LFA method to test CP from diluted rat mucosa samples (Fig. 5C). The diseased samples (red, n = 3 rats) resulted in a significantly higher (1.5 times, p< 0.05, t-test) optical density than the control samples (n = 3 rats). This equated to a higher concentration of CP in the diseased (>10 ng/ml) compared to control (0 ng/ml) samples, clearly demonstrating that the LFA format can differentiate samples obtained prior to and after the induction of colitis. As a comparison, diluted diseased and control samples were also tested using a commercial ELISA kit (Fig. 5D), and comparable results were observed, demonstrating the exceptional biosensing performance of the LFA-assisted CRISPR/Cas12a biosensor.

**Figure 5:**
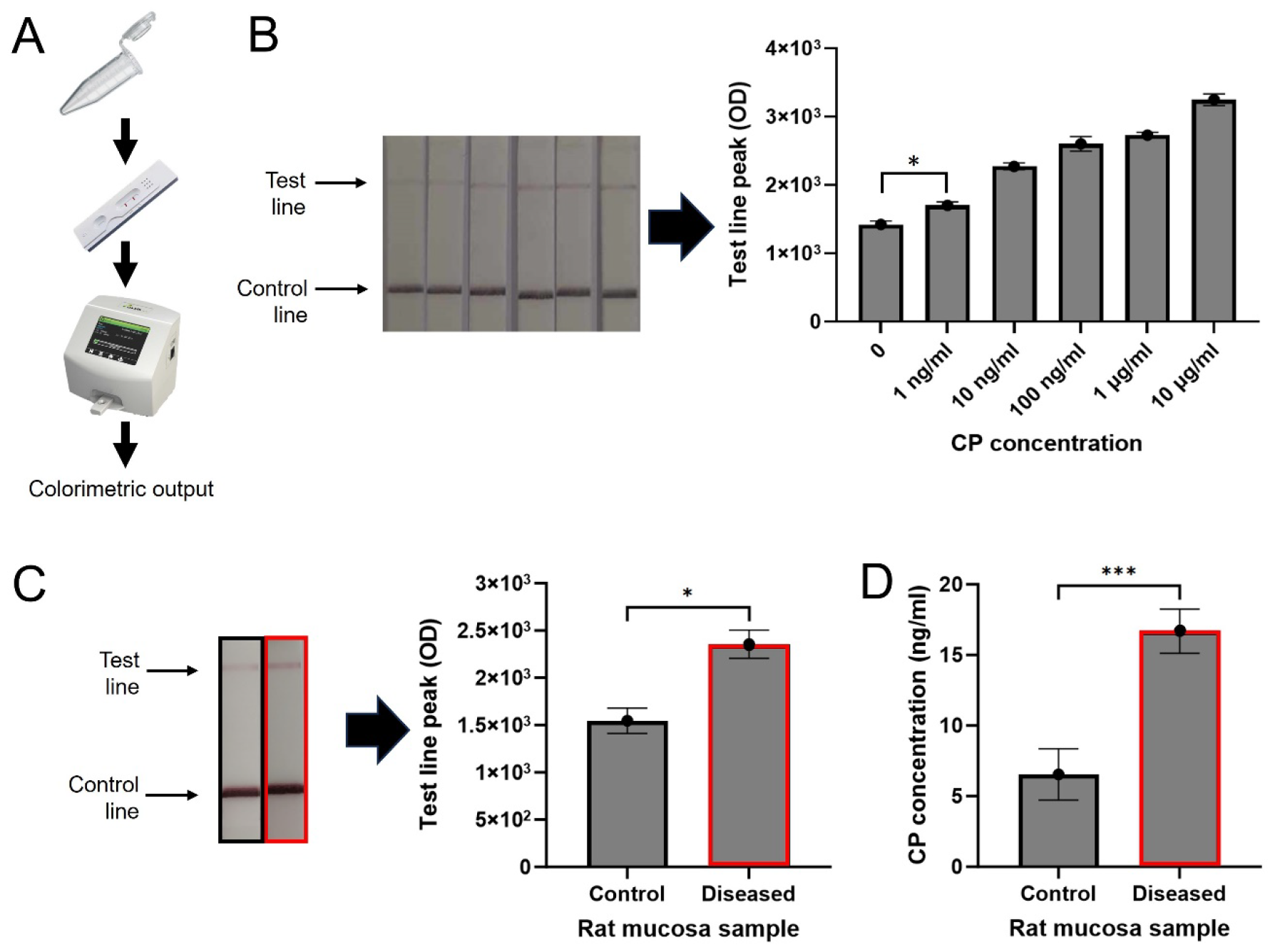
Application of LFA assisted CRISPR/Cas12a biosensor for the detection of calprotectin from rat samples. (A) Schematic of POST testing process: The CRISPR solution was mixed and pipetted into an Eppendorf tube, an LFA strip was placed into the tube, the test and control line were measured using a strip reader to get a colorimetric output of the calprotectin level; (B) Sensitivity of the LFA-assisted CRISPR/Cas12a biosensor to rat mucosa samples (n=3 repeats for each concentration). One-way ANOVA multiple comparisons, and significant differences were observed between 0 and 1 ng/mL (p<0.005). (C) Comparison of diseased rat mucosa samples (red) and the control samples (black) for a 60 min incubation time of the CRISPR buffer (n=3 rats); (D) ELISA output of diseased rat mucosa samples (red) and control samples (black) (n=3 repeats). (* *p*<0.05; ** *p*<0.01; \*\*\**p*<0.005; n=3).

## 4 Discussion

In this study, we present the potential of our CRISPR/Cas assay for the POCT of CP and demonstrated its biosensing performance in biological samples obtained from animals induced with UC. The fluorescent assay developed in this study showed a LOD of 1 ng/mL, and a detection range that spans over 4 orders of magnitude to 10 μg/mL, covering all biological concentrations of CP in the human body^35^. In addition, the fluorescent assay was successful in monitoring the CP concentration in a rat acute colitis model. To further enhance the assay and facilitate its implementation in a point-of-care setting, an LFA-assisted assay was established, which showed the same LOD and detection range as the fluorescent version. This colorimetric assay was able to differentiate healthy and diseased rat mucosa samples when only using a 60 min incubation time, underscoring its potential for point-of-care clinical applications. Furthermore, the chemical enhancer DTT, was able to enhance the performance of both assays by improving the signal strength, in line with previous work^18^.

Present CP detection technologies suffer from poor patient accessibility^36^. Our CRISPR sensors, both tube-based or in a lateral flow assay format address these deficiencies. Notably, these CRISPR sensors can measure over a 4-log detection range from 1 ng/mL to 10 μg/mL. This is significantly larger than typical turbidimetric assays which have a detection range over 2-logs, and is also wider than Quantikine ELISA’s commercial kit that has a range from 1 ng/mL to 40 ng/mL ^37^. The wider detection range offered by our CRISPR sensor is sufficient to cover most biological concentrations typically found in clinical serum samples from healthy (∼200-4000 ng/mL) and IBD (∼410-125000 ng/mL) patients^9, 10^, although mucosal CP levels have never been measured in humans previously. Additionally, the ability to differentiate healthy and diseased rat mucosa samples in an LFA format confirms the utility of our assay for POCT. This presents advantages over lab-based ELISA tests, as a rapid, accessible diagnosis method capable of producing accurate results. In addition, comparing with other established CRISPR sensors^19, 23^, our CRISPR sensor’s premise is based on CP’s enhancement effect on the output signal.

The impact of our CRISPR biosensor for IBD management is substantial when compared with conventional diagnostic approaches. The current standard for diagnosis can involve a highly invasive, costly endoscopy, or biases stemming from patients’ perception of scoring parameters utilized in multiparameter scoring and interobserver variability^5, 38, 39^. In contrast, our system offers non-invasive analysis of bodily fluid samples, providing results in 60 minutes and meets the criteria for POCT^34^, in that the sample can be analyzed near the patient and can be operated by personnel with no specialized training^40^. This enables timely results, reducing the need for specialized personnel and enhances accessibility, particularly in underserved areas ^41^. POCT also opens the potential for home testing, shifting chronic disease management away from healthcare facilities^41^. The ability to monitor disease progress over time can be achieved in an easy and convenient manner, which significantly enhances patient satisfaction and adherence to treatment plans^42, 43^. Additionally, the biosensor’s validation of disease activity through CP concentration offers the ability to establish standard diagnostic protocols, which reduces the effect of patient symptom skewness and variability between healthcare providers^42^.

A limitation of this study is that the exact mechanism of how CP enhanced the biosensing performance of the CRISPR/Cas12a biosensing system is currently not clear. As it is out of scope of this study, future studies may elucidate the mechanisms of action. From our results, we observe a positive correlation where an increased level of CP results in higher CRISPR activity, leading to a stronger fluorescence output, but further exploration is required. We postulate that the potential mechanism is due to the enhancement effect of CP on the CRISPR/Cas12a system. To date, diverse chemical enhancers to Cas12a RNP have been reported, like BSA^33^, L-porline^33^, and DTT^18^, whose potential enhancement mechanism was due to the conformation change of Cas12a RNP, but it requires further simulation at the molecular level.

## 5 Conclusion

Our CRISPR/Cas12a biosensing assay has demonstrated an excellent biosensing capability that can quantify CP concentration in fluid biological samples in 60 minutes, in a portable assay. Collectively, these aspects highlight the great potential of the biosensor in advancing the precision and rapidity of diagnosis and monitoring in chronic disease management. This innovative approach holds the promise of significantly improving patient outcomes and enhancing the overall healthcare experience for clinicians and patients dealing with IBD. Further research and clinical validation will be essential to realize the full potential of this technology in the field of IBD diagnosis and monitoring.

## Data Availability

All data produced in the present work are contained in the manuscript

## Acknowledgements

This study is supported by a UNSW SHARP fund and UNSW Mental Health & Addiction Theme Seed Funding.

